# Evaluation of methods for whole genome sequencing of *Enterococcus faecium* in a diagnostic laboratory

**DOI:** 10.1101/2021.11.09.21265457

**Authors:** Kathy E. Raven, Danielle Leek, Beth Blane, Sophia T. Girgis, Asha Akram, Nicholas Brown, Sharon J. Peacock

**Author notes:** Corresponding author: Kathy Raven. Joint first authors (Kathy E. Raven, Danielle Leek).

## Abstract

*Enterococcus faecium* is an important nosocomial pathogen associated with hospital transmission and outbreaks. Based on growing evidence that bacterial whole genome sequencing enhances hospital outbreak investigation of other bacterial species, our aim was to develop and evaluate methods for low volume clinical sequencing of *E. faecium*. Using a test panel of 22 *E. faecium* isolates associated previously with hospital transmission, we developed laboratory protocols for DNA extraction and library preparation, which in combination with the Illumina MiniSeq can generate sequence data within 24 hours. The final laboratory protocol took 3.5 hours and showed 98% reproducibility in producing sufficient DNA for sequencing. Repeatability and reproducibility assays based on the laboratory protocol and sequencing demonstrated 100% accuracy in assigning species, sequence type (ST) and (when present) detecting *vanA* or *vanB*, with all isolates passing the quality control metrics. Minor variation was detected in base calling of the same isolate genome when tested repeatedly due to variations in mapping and base calling, but application of a SNP cut-off (≤15 SNPs) to assign isolates to outbreak clusters showed 100% reproducibility. An evaluation of contamination showed that controls and test *E. faecium* sequence files contained <0.34% and <2.12% of fragments matching another species, respectively. Deliberate contamination experiments confirmed that this was insufficient to impact on data interpretation. Further work is required to develop informatic tools prior to implementation into clinical practice.

**Importance:** *Enterococcus faecium* is a leading cause of hospital infections, particularly in the immunocompromised, and has been shown to be associated with hospital transmissions. Whole-genome sequencing is a highly discriminatory technique that has been shown to be capable of identifying transmissions that may otherwise go undetected by conventional infection control methods. This could be a powerful adjunct to infection control, since previous studies have shown that there are numerous hospital outbreaks of *E. faecium*, which can extend over multiple wards and years. In this study, we developed and evaluated laboratory methods for low volume clinical sequencing of *E. faecium*, to allow uptake in smaller local diagnostic laboratories. We demonstrated that sequencing could be performed within 24-48 hours of the sample flagging positive. This would allow a more rapid turnaround time, compared to sending isolates to the reference laboratory, providing an opportunity for infection control to act earlier to prevent further transmission.

## Introduction

*Enterococcus faecium* is a common gut commensal and a leading cause of nosocomial infection, particularly in the immunocompromised (1). Infection caused by vancomycin-resistant *E. faecium* (VRE) is of particular concern since there are limited treatment options (2,3), as recognized by its inclusion in the WHO priority pathogens list for the research and development of new antibiotics (4). The majority of nosocomial infection is associated with hospital-adapted *E. faecium*, which are genetically distinct from commensal isolates (5). Based on multilocus sequence typing (MLST), hospital-adapted strains cluster within Clonal Complex (CC) 17, which has more recently been designated based on genome sequencing as clade A1 (5). Isolates belonging to this clade are globally disseminated and have been linked to numerous healthcare-associated outbreaks (6-8). Their enhanced biological fitness in healthcare settings has been associated with genetic adaptations that alter survival, virulence and antibiotic resistance (3).

The increasing application of whole genome sequencing to *E. faecium* has begun to elucidate patterns of spread at national and individual hospital levels (6-8). This includes transmission networks spanning multiple years and involving patient movement through numerous wards within the same hospital, which are difficult for infection control to detect based on the standard definition of same time and place (7). These studies have demonstrated that bacterial sequencing can bring greater resolution to healthcare-associated lineages and discriminate between isolates of the same lineage, overcoming this limitation of previous typing methods such as pulsed-field gel electrophoresis (PFGE) and MLST. Due to recombination involving regions containing the housekeeping genes used by MLST, this method has been shown to be imperfect for determining genetic relatedness (8,9), and PFGE has also been shown to cluster genetically distinct isolates (6). Our objective was to describe the development and evaluation of laboratory methodology for low volume throughput clinical sequencing of *E. faecium*.

## Materials and Methods

### Test panel isolates

Twenty-two *E. faecium* isolates were assembled into a test panel for the study (Table 1). These were selected from a study published previously (7), and represented isolates associated with bloodstream infection in patients at the Cambridge University Hospital NHS Foundation Trust hospital (CUH) in the United Kingdom between Nov 2006 and Dec 2012. Sixteen of these *E. faecium* were from four of six outbreaks described previously (7). The remainder were two *E. faecium* belonging to each of Clade A2 (animal-associated clade) and Clade B (community-associated clade); one *E. faecium* positive for *vanB*; and an *E. faecium* positive control (Table 1). The collection included vancomycin-resistant and -susceptible isolates.

**Table 1.** Panel of isolates used in this study

| ID | Accession number | Species | Vancomycin resistance | ST | Reason for inclusion | Repeatability | Reproducibility | Contamination |
| --- | --- | --- | --- | --- | --- | --- | --- | --- |
| EC0397 | ERR1820598 | *E. faecium* | VSE | 203 | Cluster A | Included | Included |  |
| EC0102 | ERR1820597 | *E. faecium* | *vanA* | 203 | Cluster A |  | Included |  |
| EC0333 | ERR370022 | *E. faecium* | VSE | 203 | Cluster A |  |  |  |
| EC0503 | ERR1820599 | *E. faecium* | *vanA* | 203 | Cluster A |  | Included |  |
| EC0130 | ERR375305 | *E. faecium* | *vanA* | 375 | Cluster B |  | Included |  |
| EC0518 | ERR388722 | *E. faecium* | *vanA* | 375 | Cluster B | Included | Included |  |
| EC0185 | ERR377443 | *E. faecium* | *vanA* | 375 | Cluster B | Included |  |  |
| EC0163 | ERR375422 | *E. faecium* | *vanA* | 375 | Cluster B | Included | Included |  |
| EC0129 | ERR370001 | *E. faecium* | *vanA* | 117 | Cluster C |  | Included |  |
| EC0175 | ERR375395 | *E. faecium* | *vanA* | 117 | Cluster C |  |  |  |
| EC0142 | ERR375299 | *E. faecium* | *vanA* | 117 | Cluster C |  | Included |  |
| EC0146 | ERR375367 | *E. faecium* | *vanA* | 117 | Cluster C |  | Included |  |
| EC0031 | ERR370024 | *E. faecium* | *vanA* | 132 | Cluster D |  | Included |  |
| EC0028 | ERR377427 | *E. faecium* | *vanA* | 132 | Cluster D |  | Included |  |
| EC0036 | ERR369956 | *E. faecium* | *vanA* | 132 | Cluster D |  | Included |  |
| EC0046 | ERR375456 | *E. faecium* | *vanA* | 132 | Cluster D |  |  |  |
| EC0181 | ERR375360 | *E. faecium* | *vanB* | 203 | *vanB* positive | Included |  |  |
| EC0333 | ERR370022 | *E. faecium* | VSE | 203 | Clade A2 |  |  |  |
| EC0322 | ERR375339 | *E. faecium* | VSE | Novel | Clade A2 |  |  |  |
| EC0214 | ERR375555 | *E. faecium* | *vanA* | 203 | Cluster A |  |  |  |
| EC0392 | ERR375541 | *E. faecium* | VSE | Novel | Clade B |  |  |  |
| EC0160 | ERR377442 | *E. faecium* | *vanA* | 203 | Positive control, Cluster A | Included | Included | Included |
| NCTC12241 | ERR718772 | *E. coli* | None | Not applicable | Negative control | Included | Included | Included |
| No template | Not applicable | None | None | None | No template control | Included | Included | Included |

Cluster names A-D indicate individual outbreaks of *E. faecium* described previously (7).

### DNA extraction, library preparation and sequencing

Isolates were cultured from frozen stocks onto CBA and incubated at 37°C for 48 hours. A single colony was then sub-cultured onto CBA, incubated at 37°C for 48 hours and frozen at −80°C in Microbank vials. For DNA extraction and subsequent sequencing, isolates were taken from these pure stocks and incubated at 37°C on CBA overnight. DNA extraction was performed manually using QIAgen kits according to the published QIAamp DNA Mini and Blood Mini Handbook protocol, following ‘Appendix D: Protocols for Bacteria – Isolation of genomic DNA from Gram-positive bacteria’ with the following amendments: (i) colonies were used direct from the culture plate instead of pelleting bacteria, (ii) the lysozyme solution was made using water and EDTA, (iii) the 95°C incubation was removed, (iv) in steps 8 and 11 the centrifuge speed was set to maximum (13,200rpm), (v) the option of 50μl of distilled water was used for elution including the 5 minute incubation step, with two 50μl volumes filtered through in succession (final volume of 100μl), and 50μl of this used for a final filter. Further amendments to the DNA extraction protocol were made as described in the results. DNA was quantified post-extraction using a Qubit fluorometer and the dsDNA HS assay kit (Thermofisher, UK). Library preparation was performed as described previously using the Illumina Nextera Flex kit (10). Sequencing was performed on an Illumina MiniSeq with a run-time of 13 hours, using the high output 150 cycle MiniSeq cartridge and Generate Fastq workflow. Data were transferred to an external 1TB USB-connected hard drive. Based on an expected total data output of 3.3-3.8Gb, an average genome size of 2.9MB (https://www.ncbi.nlm.nih.gov/genome/?term=Enterococcus%20faecium[Organism]&cmd=DetailsSearch), and a target of 50x coverage, we estimated that approximately 24 *E. faecium* isolates could be sequenced on a single run (estimated 47-55x coverage). We therefore included 21 test *E. faecium* isolates and three controls (*E. coli, E. faecium* and no template, see below) per sequence run for the initial reproducibility runs.

### Sequence data analysis

Multilocus sequence types (ST) of the *E. faecium* isolates were identified using ARIBA version 2.12.1 as described at https://github.com/sanger-pathogens/ariba/wiki/MLST-calling-with-ARIBA. Species were determined using Kraken version 1 (https://ccb.jhu.edu/software/kraken/) with the miniKraken database available at https://ccb.jhu.edu/software/kraken/dl/minikraken_20171019_8GB.tgz. The presence of *vanA* and *vanB* was determined using Ariba with *vanA* from M97297 (positions 6979-8010) and *vanB* from Aus0004 (accession number CP003351, positions 2839377-2840405) as reference genes. All isolates were mapped to the *E. faecium* strain Aus0004 (accession number CP003351) using SMALT (https://www.sanger.ac.uk/science/tools/smalt-0) with mapping and base calling performed as described previously (11), with the following modifications: kmer size 13, step size 6. The depth and percentage coverage of the mapping reference were determined using the script available at https://github.com/sanger-pathogens/vr-codebase/blob/master/lib/VertRes/Pipelines/Mapping.pm. Mobile genetic elements (MGEs) were removed using the script available at https://github.com/sanger-pathogens/remove_blocks_from_aln. SNPs were identified based on the following parameters: minimum number of reads matching the SNP = 4; minimum number of reads matching the SNP per strand = 2; ratio of SNP base to alternative base >0.75; variant quality >50; mapping quality >30.

SNP distances between isolate pairs reported previously (7) were defined after removal of recombination regions using Gubbins across a large collection of clade A1 isolates. We elected not to remove recombination in this study because looking to the future when genomes are analysed using fully automated tools, recombination removal will currently be challenging to incorporate into tools that offer very rapid interpretation. To correct for this during the reproducibility of the pairwise SNP distance with the original sequence dataset, we recalculated this based on mapping and removal of MGEs, but without regions of recombination removed. These updated values were used as the ‘expected’ number of SNPs between isolates for the repeatability and reproducibility runs.

### Positive and negative controls

Three controls were included in every sequencing run to monitor the ongoing performance of the entire testing process. These were a no template control, a positive control (*E. faecium* EC0160), and a negative control (*E. coli* NCTC12241). We selected a positive control that was a known number of core genome SNPs (n=31) from another isolate sequence (EC0037) from the same collection, to control for base calling in the place of PhiX. The positive control was used to control the entire assay process and analytical accuracy. The negative control was used to assess cross-contamination during processing and represented the non-target DNA sample to verify analytical specificity. Fresh stocks of molecular grade water and phosphate-buffered-saline were opened each week. The no template control contained all assay components except for DNA and was used to verify the lack of contamination across reagents and samples. Other ‘reuse’ reagents were checked for bacterial contamination weekly by sub-culturing using a 10µl loop onto CBA and incubating for 48 hours in air at 37°C.

### Sequence metrics for controls

Controls were required to pass the following quality metrics. *E. faecium* positive control: highest match to *E. faecium* using Kraken, assigned to ST203, *vanA* detected, minimum mean sequence depth of 20x and minimum 80% coverage of the mapping reference genome (Aus0004). *E. coli* negative control: highest species match to *E. coli* in Kraken, *vanA* not detected, no *E. faecium* ST assigned. No template control: contamination from any bacterial DNA of less than 30,000 fragments in Kraken. *E. faecium* isolates from the test panel were required to pass the following metrics: highest match to *E. faecium* using Kraken, assigned to the correct ST, *vanA* detected or not detected as appropriate (Table 1), minimum sequence depth of 20x and minimum 70% coverage of the mapping reference genome. This lower value of 70% coverage was used for test isolates since Clade A2 and Clade B isolates are more distantly related to the Clade A1 reference genome. Analysis of 799 genomes reported previously (7,8) identified 17 genomes with <80% coverage of Aus0004 (74.8-79.6% coverage), which belonged to Clade B (n=10), Clade A2 (n=4) or Bayesian Analysis of Population Structure (BAPS) group 5 (n=3).

### Repeatability and reproducibility

Repeatability was evaluated by sequencing six *E. faecium* isolates (EC0160 (positive control), EC0102, EC0181, EC0333, EC0397 and EC0503 (Table 1)) in triplicate in a single sequencing run. Reproducibility was evaluated by sequencing 21 *E. faecium* isolates from the test panel in three independent runs, and subsequently repeated with 12 *E. faecium* isolates in three independent runs (see results). For each isolate, the pure single-colony frozen stock was sub-cultured onto three separate CBA plates and incubated in air at 37°C overnight, two heaped 1μl loops (Supplementary Figure 1) from each of these plates were then taken forwards for individual DNA extraction, library preparation and sequencing. The entire process for the reproducibility experiments was performed by different laboratory staff on three different days. The resulting fastq files were analysed as above.

Isolates were classified as part of the same cluster if they were ≤15 SNPs apart. This cut-off was selected based on a mutation rate of 7 SNPs/genome/year (7) and a within-host diversity of 6 SNPs (12,13), using the formula described previously (14) to capture transmission within 6 months. Isolates >15 SNPs different were classified as genetically unrelated. Sensitivity and specificity for allocation of isolates into outbreaks were calculated using the following definitions: true positives, the number of genetically related isolates based on the original data that cluster together based on the test data; false negatives, the number of genetically related isolates based on the original data that do not cluster together in the test data; true negatives, the number of genetically unrelated isolates based on the original data that do not cluster together in the test data; and false positives, the number of genetically distant isolates based on the original data that cluster together based on the test data (15).

### Analysis of contamination

The impact on quality metrics from varying levels of DNA contamination during clinical *E. faecium* sequencing was evaluated using intentional spiking experiments. The *E. faecium* positive control (EC0160), the *E. coli* negative control (NCTC 12241), and an *E. faecalis* isolate (NCTC 13779) (selected because *E. faecalis* are commonly found on clinical plates with *E. faecium*) were cultured and DNA extracted and quantified as described above. Donor (contaminating) DNA was inoculated into the recipient (true) sample to achieve a final spiked concentration of 0%, 0.1%, 1%, 10% or 20%. The donor-recipient combinations were as follows: (i) recipient EC0160, donor NCTC 12241; (ii) recipient EC0160, donor NCTC 13779; (iii) recipient water, donor EC0160. Contamination with the spike was defined based on the number and proportion of fragments matching to *E. faecium, E. faecalis* or *E. coli* based on Kraken. The effect of contamination was evaluated using this metric together with the proportion of the *E. faecium* reference covered during mapping, depth of coverage of the mapping reference, and *vanA* and ST detected by Ariba. Additional unintentional contamination from internal controls or external sources was evaluated based on the number and proportion of reads matching to other species in Kraken.

### Data availability

Sequence data generated during this study are available from the European Nucleotide Archive (https://www.ebi.ac.uk/ena) under the accession numbers listed in Table 1.

## Results

We sought to develop and describe methods for low-throughput *E. faecium* sequencing in a routine microbiology laboratory within a 24-hour turnaround time (from DNA extraction to availability of sequence data). This included an evaluation of quality controls, precision (reproducibility and repeatability), and contamination.

First, we determined whether it was possible to extract DNA from a colony picked from the primary clinical culture plate. Following the default DNA extraction protocol, we found that extraction from a single colony after either 24 hours or 48 hours of incubation did not provide enough DNA for input to the library preparation protocol, defined as being less than the 3.3ng/μl required (Supplementary Table 1). We concluded that using colonies from the primary plate was not feasible, and sub-cultured a single colony onto CBA and incubated for a further 24 hours to create purity plates. DNA extraction from these purity plates demonstrated that a single heaped 1μl loop input (Supplementary Figure 1) produced insufficient DNA in 10% of cases (Supplementary Table 1), whilst two heaped 1μl loops provided the required amount of DNA in all cases (Supplementary Table 1). We therefore proceeded with two 1μl heaped loops input to DNA extraction.

We next aimed to determine whether the DNA extraction protocol time could be reduced from the current time of 2 hours. Comparison of the DNA output using 30-minute incubations for proteinase K and buffer AL versus 15-minute incubations at these steps revealed that both methods resulted in sufficient DNA (Supplementary Table 1). We therefore proceeded with a final protocol of two heaped 1μl loops input and 15 minutes incubation for proteinase K and buffer AL, which reduced the time for DNA extraction from 2 to 1.5 hours. The final DNA extraction protocol was performed three times by three different people, which demonstrated 98% reproducibility (65/66) for acquiring sufficient DNA for input to library preparation (Supplementary Table 1).

Using the shortened DNA extraction protocol and previously described reduced library preparation protocol (10), we aimed to determine the repeatability and reproducibility of the full sequencing protocol. Repeatability was based on concordance of assay results and quality metrics for six *E. faecium* isolates sequenced in triplicate in a single sequencing run. There was 100% concordance in assigning species, ST and detecting *vanA* and *vanB* (Supplementary Table 2). Analysis of the pairwise SNP differences between the within-run replicates found that four of the six isolates were genetically identical across all three replicates, whilst the remaining two isolates (EC0397 and EC0503) had 0-2 and 0-5 SNPs, respectively, different between the three replicates. In the case of EC0397, replicates 1 and 3 were identical, but replicate 2 differed by 1-2 SNPs, whilst for EC0503 replicates 1 and 2 were identical but replicate 3 differed by 4-5 SNPs. This provided a repeatability per replicate of 78% (14/18) based on a requirement for isolates to be identical, increasing to a repeatability per replicate of 89% (16/18) if small variations in SNPs (≤2 SNPs) were allowed. Analysis of the sequence files of EC0397 revealed that there were three positions where the base calls varied between replicates. At two of these positions, both bases were detected but in different proportions, while the third variable position could be explained by misalignment around an indel. By contrast, EC0503 was identical in run 1 and run 2, but 5 SNPs different in run 3. Four of the five SNPs were located in a single 5bp region and were likely caused by misalignment around an indel. Using the original published sequence mapped to the Aus0004 reference with MGEs removed as the ‘expected’ number of SNPs, the 6 isolates in triplicate had between 0-6 base calls different to the original sequence (excluding positions denoted as ‘N’ because of failure to call a base), which is within the expected within-patient diversity of 6 SNPs (Supplementary Table 2).

We initially evaluated reproducibility by sequencing 21 test panel *E. faecium* isolates in three independent sequence runs (Supplementary Table 2). However, the average depth of coverage across the three runs was lower than expected (43x, range 16.4-72.1x) despite a higher data output than expected (4-5.2Gb compared to an expected 3.3-3.8Gb). Calculation of the estimated genome size based on the data output and average depth of coverage across the three sequence runs indicated a genome size of ∼4.8Mb (Supplementary Table 3). Investigation of the mapping files revealed that this could be caused by high depth of coverage of plasmid elements, possibly due to multiple plasmid copies. Based on an expected data requirement of ∼4.8Mb, we estimated that 12 isolates plus three controls per sequencing run would produce sufficient data to obtain 50x coverage. Repeats of the reproducibility experiments with 12 isolates and 3 controls revealed an average depth of coverage of 88.6x, with a range of 43.2-113.6x (Supplementary Table 2) based on 5.4-6.5Gb data output, compared to an expected 89.9x coverage based on the observed data output.

Across the reproducibility runs with 12 isolates and 3 controls there was 100% accuracy in assigning species, ST and detecting *vanA*. There were 0-3 SNPs identified for between-run replicates, providing a reproducibility per replicate of 67% (26/39) based on replicates needing to be identical, and a reproducibility per replicate of 92% (37/39) if small variations (≤2 SNPs) were allowed between runs. Two isolates (EC0130 and EC0142) had 3 SNPs different between runs. Analysis of the genomes revealed that in EC0130 these SNPs were clustered in a 60bp region where a short section of reads had mapped in a region with poor or no mapping, and both bases were present in the read files in all three repeats. In EC0142 two SNPs were adjacent in the genome in a short intergenic region lacking mapped reads in the bam file, whilst the third was located in a short poorly mapped region in an otherwise absent gene where all three repeats had a mixture of base calls at the position. Using the original published sequence when mapped to the Aus0004 reference with MGEs removed as the ‘expected’ number of SNPs, the new sequences were 0-4 SNPs different, which is within the expected within-host diversity (6 SNPs).

We next sought to determine the sensitivity and specificity for outbreak detection in each of the three reproducibility runs, using the genetic relatedness established previously as the gold standard. The 12 isolates represented four distinct outbreaks encompassing four different STs (ST203, ST375, ST117 and ST132) identified during 6 years of genomic surveillance. All 12 isolates (36 isolate pairs) were classed in the same relatedness category (0-15 SNPs, >15 SNPs) in each of the three sequence runs. This provides a sensitivity and specificity for outbreak detection between runs of 100%. However, when the reproducibility data was compared to the original sequence data, there were 2/12 discrepancies in the classification of isolate pairs (one pair from Cluster A, one pair from Cluster D). Both pairs were called as related by the original data (7 SNPs) and unrelated by the new data (16-24 SNPs). Analysis of the SNP locations revealed that the majority of the discrepant SNPs (10/11 and 21/23 discrepant SNP locations in Cluster D and A, respectively) were due to a single poorly mapped region which contained bacteriocin and plasmid genes and was located between two ISEfm transposases, which had been called as unknown bases in the original sequence. It was also found that 4 of the 7 SNPs in the original sequence data between the isolate pair in Cluster A were clustered in a single 72bp region with poor mapping.

To determine the impact of DNA contamination (for example from cross-contamination during processing) we performed deliberate contamination experiments. Details of the donor and recipient DNA, the concentrations of spiked DNA and our findings are summarized in Table 2. Contamination of the no template control with increasing concentrations of *E. faecium* DNA did not lead to the control erroneously passing the QC metrics for *E. faecium* until the final spiked concentration reached >10%. This indicates that contamination of the no template control at 1% (which equated to 30,452 fragments matching *E. faecium* in Kraken) can be tolerated. Contaminating the positive *E. faecium* control with increasing concentrations of *E. coli* and *E. faecalis* DNA demonstrated that this could tolerate up to 10% contamination (which equated to 8.92 or 11.34% fragments, respectively, in Kraken) before the *E. faecium* QC metrics were not achieved. Contamination with up to 20% *E. coli* or *E. faecalis* did not appear to affect SNP calling (Supplementary Table 4).

**Table 2.** Results of deliberate contamination experiments

| Question | Recipient | Donor | Concentration of contamination | Number/proportion of reads matching donor species | Proportion of reads matching *E. faecium* | Coverage of mapping reference | Average depth | MLST | *van* | QC metrics passed for positive control |
| --- | --- | --- | --- | --- | --- | --- | --- | --- | --- | --- |
| Determine the effect of contaminating the no template control with increasing concentrations of *E. faecium* DNA | No template | *E. faecium* | 0% | 317 | n/a | 1.1 | 0.0 | ND | None | No |
|  |  |  | 0.1% | 1725 | n/a | 6.1 | 0.1 | ND | None | No |
|  |  |  | 1% | 30,452 | n/a | 60.0 | 1.1 | ND | Partial match | No |
|  |  |  | 10% | 42,015 | n/a | 86.9 | 11.4 | 203 | *vanA* | No |
|  |  |  | 20% | 47,621 | n/a | 87.4 | 20.6 | 203 | *vanA* | Yes |
| Determine the effect of contaminating the *E. faecium* control with increasing concentrations of another organism | *E. faecium* control | *E. coli* control | 0% | 0.01% | 85.95% | 88.2 | 53.0 | 203 | *vanA* | Yes |
|  |  |  | 0.1% | 0.14% | 85.73% | 88.2 | 53.4 | 203 | *vanA* | Yes |
|  |  |  | 1% | 0.99% | 84.05% | 87.9 | 42.1 | 203 | *vanA* | Yes |
|  |  |  | 10% | 8.92% | 72.29% | 88.2 | 49.7 | 203 | *vanA* | Yes |
|  |  |  | 20% | 16.43% | 61.12% | 88.3 | 50.0 | 203 | *vanA* | Yes |
|  | *E. faecium* control | *E. faecalis* | 0% | 0.18% | 85.86% | 88.0 | 47.2 | 203 | *vanA* | Yes |
|  |  |  | 0.1% | 0.35% | 85.75% | 87.9 | 44.0 | 203 | *vanA* | Yes |
|  |  |  | 1% | 1.37% | 84.75% | 88.3 | 54.0 | 203 | *vanA* | Yes |
|  |  |  | 10% | 11.34% | 74.84% | 88.5 | 43.5 | 203* | *vanA* | No |
|  |  |  | 20% | 21.92% | 64.12% | 88.5 | 37.3 | 203* | *vanA* | No |

\* indicates that there was uncertainty in the MLST call by Ariba.

We also evaluated unintentional contamination in the seven runs (excluding the deliberate contamination assay). The maximum proportion of fragments matching another species was 0.34% for the controls and 2.12% for the test isolates, with the highest matches being those of related species such as *Enterococcus durans* and *Lactococcus lactis*. Based on the number of fragments in Kraken for the no template controls and the proportion of fragments in Kraken for the remaining sequences, this demonstrates that all controls and test isolates had levels of contamination below 1% (Supplementary Table 2).

## Discussion

We aimed to develop manual methods for low-throughput sequencing of clinical *E. faecium*. This would allow uptake of clinical sequencing in smaller local diagnostic laboratories that lack the capacity for high-throughput sequencing. At present, isolates suspected to be part of an outbreak are sent to the public health reference laboratory for further testing, with results taking approximately two weeks to be returned (16). However, as the cost of sequencing and sequencers reduces, the possibility of in-house sequencing at local clinical laboratories will become more viable. This will allow a more rapid turnaround time to detection of outbreaks, providing the opportunity for infection control to act earlier to prevent further transmission. This is important since previous studies have shown that there are multiple outbreaks in a hospital based on bloodstream infections alone, which represent only the tip of the iceberg for transmission (7). Here we have shown that sequencing can be performed within 24-48 hours of a sample flagging as positive, providing a rapid tool to aid infection control.

In the clinical laboratory, two important factors for consideration are turnaround time and cost. We aimed to identify the shortest turnaround-time from clinical plates to sequence data. We found that single colonies produced insufficient DNA for sequencing, likely due to the small colony size, meaning that purity plates are required. Purity plates may already be available in the clinical laboratory at the time the sample flags as positive (for example, purity plates used for disc testing), allowing a 24 hour turnaround time, whilst the remainder will require a 48 hour turnaround time. This still represents a faster turnaround time than testing at the reference laboratory. We were also able to reduce the hands-on processing time from 4 hours to 3.5 hours for DNA extraction and library preparation. Since library preparation does not require normalization (a tricky and potentially time-consuming step), this represents a rapid and simple method for sequencing. To reduce the cost of sequencing per sample we aimed to maximise the number of *E. faecium* isolates per sequence run. We found that 21 isolates and 3 controls passed the quality control metrics in 95% of cases and had a lower than expected coverage depth, potentially due to multiple copy number plasmids increasing the data requirements for each isolate. We therefore proceeded with 12 isolates and 3 controls.

Whilst the sequence data had 100% repeatability and reproducibility for assigning species, ST and *van* genes, SNP detection was more variable. The majority of variation within and between runs was minimal (0-1 SNPs) leading to a repeatability of 89% and reproducibility of 92% if small variations (≤2 SNPs) were allowed, and the sensitivity and specificity for outbreak detection between runs was 100%. However, some isolates had up to 5 SNPs different within and between runs, and the sensitivity for outbreak detection dropped to 67% when compared to the original sequence data. These discrepancies were explained by issues with mapping and variant calling such as misalignment around indels and mapping of short segments of the genome with high-density SNPs, including regions associated with mobile genetic elements. Options for reducing these SNP errors could be to include recombination detection, although this would be challenging to implement in rapid automated analysis tools; improve the MGE file; use a core genome scheme such as that suggested by Coll et al. (14), although this will need to be tailored to Clade A1 since community isolates are considered genetically distant enough to classify as a different species (17); or alter the mapping quality filters to reduce the impact of poorly mapped regions. The latter has been used previously to improve detection of heterozygous sites in MRSA, where sites are only considered if they are >50bp apart. Further work is needed to determine the optimum SNP analysis methodology for this species.

Finally, we determined the level of contamination that could be tolerated and evaluated the quality control metrics. All controls, test panel isolates and clinical isolates passed the required quality control metrics. We found through a deliberate contamination experiment that the data could be contaminated by up to 10% before quality control metrics were failed, whilst all test isolates and controls showed <1% contamination.

Our findings indicate that the methods evaluated here can provide high quality sequence data, and represents the first step towards being able to perform real-time clinical sequencing of *E. faecium*. However, further work is required to develop data interpretation software that can resolve issues relating to SNP calling in the *E. faecium* genome. This will be required before rapid automated analysis tools can be developed for easy interpretation by clinical staff.

## Acknowledgements

This publication presents independent research supported by the Health Innovation Challenge Fund (WT098600, HICF-T5-342), a parallel funding partnership between the Department of Health and Wellcome Trust. The views expressed in this publication are those of the author(s) and not necessarily those of the Department of Health or Wellcome Trust.

---

**Supplementary figure 1:**
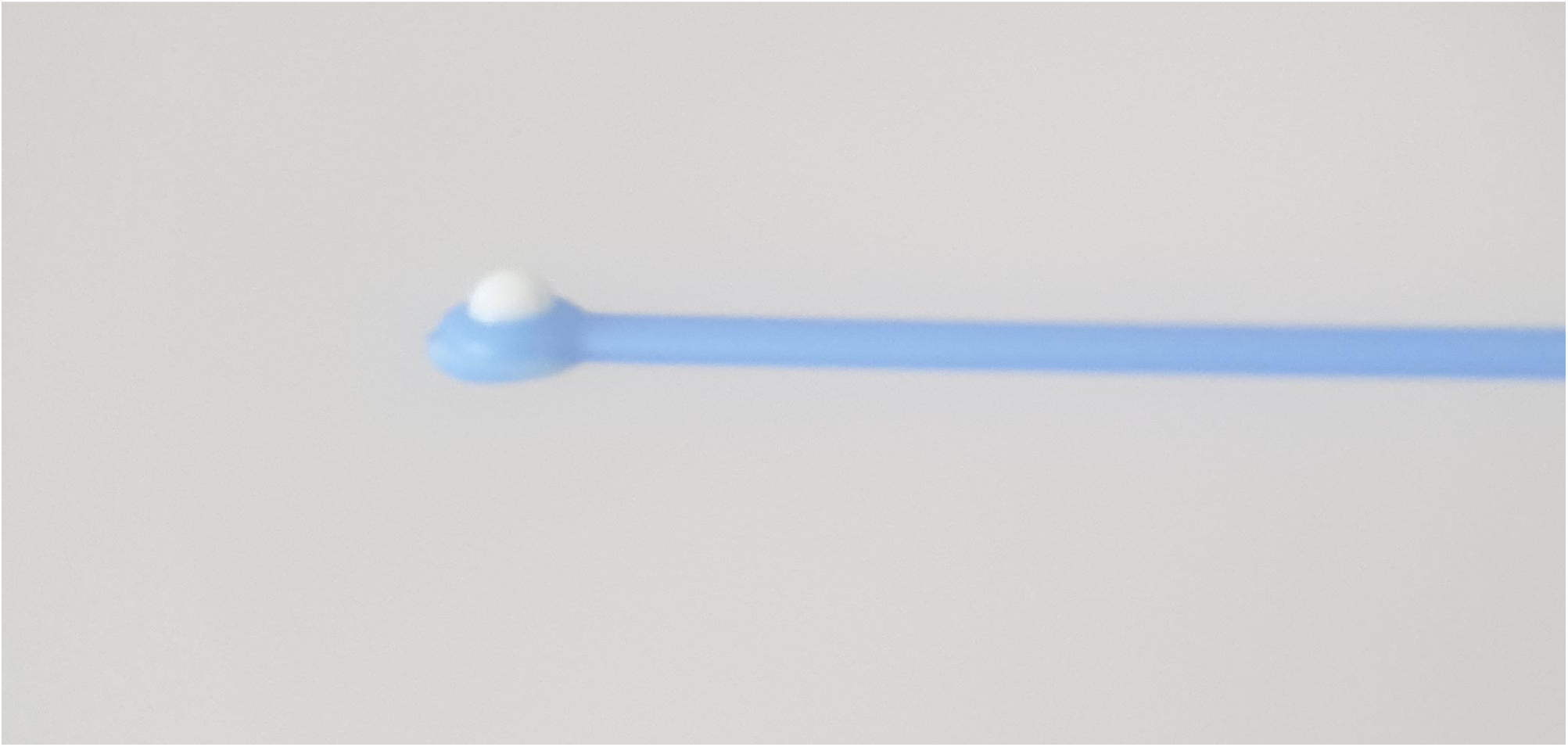
Figure showing a heaped 1μl loop of E. faecium used as input to DNA extraction

## References

1. Arias CA, Murray BE. The rise of the Enterococcus: beyond vancomycin resistance. Nat Rev Microbiol 2012. 10(4): 266–278.

2. Werner G, Coque TM, Hammerum AM, Hope R, Hryniewicz W, Johnson A, Klare I, Kristinsson KG, Leclercq R, Lester CH, Lillie M, Novais C, Olsson-Liljequist B, Peixe LV, Sodowy E, Simonsen GS, Top J, Vuopio-Varkila J, Willems RJ, Witte W, Woodford N. Emergence and spread of vancomycin resistance among enterococci in Europe. Eurosurveillance 2008. 13(47), 19046.

3. Cattoir V, Leclercq R. Twenty-five years of shared life with vancomycin-resistant enterococci: is it time to divorce? J Antimicrob Chemother. 2013. 68(4): 731–742.

4. World Health Organisation. Available online: https://www.who.int/news/item/27-02-2017-who-publishes-list-of-bacteria-for-which-new-antibiotics-are-urgently-needed (accessed on 11/06/2021).

5. Lebreton F, van Schaik W, McGruire AM, Godfrey P, Griggs A, Mazumdar V, Corander J, Cheng L, Saif S, Young S, Zeng Q, Wortman J, Birren B, Willems RJL, Earl AM, Gilmore MS. Emergence of epidemic multidrug-resistant Enterococcus faecium from animal and commensal strains. mBio 2013. 20;4(4):e00534–13.

6. Pinholt M, Larner-Svensson H, Littauer P, Moser CE, Pederson M, Lemming LE, Ejlertsen T, Sondergaard TS, Holzknecht BJ, Justesen US, Dzajic E, Olsen SS, Nielsen JB, Worning P, Hammerum AM, Westh H, Jakobsen L. Multiple hospital outbreaks of vanA Enterococcus faecium in Denmark, 2012-13, investigated by WGS, MLST and PFGE. J Antimicrob Chemother 2015. 70(9): 2474–82.

7. Raven KE, Gouliouris T, Brodrick H, Coll F, Brown NM, Reynolds R, Reuter S, Torok ME, Parkhill J, Peacock SJ. Complex routes of nosocomial vancomycin-resistant Enterococcus faecium transmission revealed by genome sequencing. Clin Infect Dis 2017. 64(7):886–893.

8. Raven KE, Reuter S, Reynolds R, Brodrick HJ, Russell JE, Torok ME, Parkhill J, Peacock SJ. A decade of genomic history for healthcare-associated Enterococcus faecium in the United Kingdom and Ireland. Genome Res 2016. 26(10)|: 1388–1396.

9. Howden BP, Holt KE, Lam MMC, Seemann T, Ballard S, Coombs GW, Tong SYC, Grayson ML, Johnson PDR, Stinear TP. Genomic insights to control the emergence of vancomycin-resistant enterococci. mBio. 2013. 4(4): e00412–13.

10. Raven KE, Blane B, Leek D, Churcher C, Kokko-Gonzales P, Pugazhendhi D, Fraser L, Betley J, Parkhill J, Peacock SJ. Methodology for whole-genome sequencing of methicillin-resistant Staphylococcus aureus isolates in a routine hospital microbiology laboratory. J Clin Microbiol. 2019. 57(6):e00180–19.

11. Klemm EJ, Shakoor S, Page AJ, Qamar FN, Judge K, Saeed DK, Wong VK, Dallman TJ, Nair S, Baker S, Shaheen G, Qureshi S, Yousafzai MT, Saleem MK, Hasan Z, Dougan G, Hasan R. Emergence of an extensively drug-resistant Salmonella enterica serovar typhi clone harboring a promiscuous plasmid encoding resistance to fluoroquinolones and third-generation cephalosporins. mBio 2018. 9(1): e00105–18.

12. Gouliouris T, Coll F, Ludden C, Blane B, Raven KE, Naydenova P, Crawley C, Torok ME, Enoch DA, Brown NM, Harrison EM, Parkhill J, Peacock SJ. Quantifying acquisition and transmission of Enterococcus faecium using genomic surveillance. Nat Microbiol 2021. 6(1):103–111.

13. Brodrick HJ, Raven KE, Harrison EM, Blane B, Reuter S, Torok ME, Parkhill J, Peacock SJ. Whole-genome sequencing reveals transmission of vancomycin-resistant Enterococcus faecium in a healthcare network. Genome Med. 2016 8(1): 4.

14. Coll F, Raven KE, Knight GM, Blane B, Harrison EM, Leek D, Enoch DA, Brown NM, Parkhill J, Peacock SJ. Definition of a genetic relatedness cutoff to exclude recent transmission of methicillin-resistant Staphylococcus aureus: a genomic epidemiology analysis. Lancet Microbe 2020. 1(8): e328–335.

15. Kozyreva VK, Truong CH, Greninger A, Crandall J, Mukhopadhyay RC, Chaturvedi V. Validation and implementation of clinical laboratory improvements act-compliant whole-genome sequencing in the public health microbiology laboratory. J Clin Microbiol 2017. 55(8):2502–2520

16. National Infection Service. Bacteriology Reference Department User Manual. Version 13, October 2020. Available online: https://assets.publishing.service.gov.uk/government/uploads/system/uploads/attachment_data/file/926234/BRDW0078.13_BRD_User_Manual.pdf (accessed on 17/06/2021)

17. Palmer KL, Godfrey P, Griggs A, Kos VN, Zucker J, Desjardins C, Cerqueira G, Gevers D, Walker S, Wortman J, Feldgarden M, Haas B, Birren B, Gilmore MS. Comparative genomics of enterococci: variation in Enterococcus faecalis, clade structure in E. faecium, and defining characteristics of E. gallinarum and E. casseliflavus. mBio 2012 3(1): e00318–11.

